# Normalization of Cerebral Blood Flow, Neurochemicals, and White Matter Integrity After Kidney Transplantation

**DOI:** 10.1101/2020.05.04.20091199

**Authors:** Rebecca J. Lepping, Robert N. Montgomery, Palash Sharma, Jonathan D. Mahnken, Eric D. Vidoni, In-Young Choi, Mark J. Sarnak, William M. Brooks, Jeffrey M. Burns, Aditi Gupta

## Abstract

**Background:** Chronic kidney disease (CKD) is associated with abnormalities in cerebral blood flow (CBF), cerebral neurochemical concentrations and white matter integrity, each of which are associated with adverse clinical consequences in the non-CKD population, and may explain the high prevalence of dementia and stroke in end stage kidney disease (ESKD). Since cognition improves after kidney transplantation (KT), we examined these brain abnormalities pre-to post-KT to identify potential reversibility in ESKD-associated brain abnormalities.

**Methods:** We measured the effects of KT on CBF assessed by arterial spin labeling, cerebral neurochemical concentrations (N-acetylaspartate, choline, glutamate and glutamine, myoinositol and total creatine) measured by magnetic resonance spectroscopic imaging, and white matter integrity measured by fractional anisotropy (FA) and mean diffusivity (MD) with diffusion tensor imaging. We used a linear mixed model analysis to compare longitudinal, repeated brain MRI measurements pre-KT, and 3 months and 12 months post-KT, and also compared findings with healthy controls.

**Results:** 29 ESKD patients and 19 age-matched healthy controls participated in the study. 22 patients underwent post-KT MRI. CBF, which was higher pre-KT than in controls (p=0.003), decreased post-KT (p<0.0001) to values in controls. KT also normalized concentrations of osmotic neurochemicals choline (p<0.0001) and myo-inositol (p=0.0003) that were higher pre-KT compared to controls. Post-KT, FA increased (p=0.001) and MD decreased (p=0.0001).

**Conclusions:** Brain abnormalities in CKD are reversible and normalize with KT. Further studies are needed to understand the mechanisms underlying these brain abnormalities and to explore interventions to mitigate them even in patients who cannot be transplanted.

**Significance statement:** Kidney disease is accompanied by brain structural and physiological abnormalities and increased risk of dementia and stroke. Renal replacement therapy with dialysis does not normalize these brain abnormalities. We evaluated these brain abnormalities before and after kidney transplantation and demonstrated that unlike dialysis, kidney transplantation normalizes cerebral blood flow, neurochemical concentrations and white matter integrity. These changes persist beyond initial post-transplantation period and thus cannot be attributed to peri-procedural interventions like steroids. These results indicate reversibility of brain abnormalities in kidney disease. Further studies are needed to understand the mechanisms underlying these abnormalities and explore interventions for prevention and mitigation in patients who cannot be transplanted.

## Introduction

Chronic kidney disease (CKD) is associated with increased risk of dementia, stroke and mortality.^1^ For patients with end stage kidney disease (ESKD), dialysis is a lifesaving procedure, but initiation of dialysis further increases strokes and mortality.^2^ These deleterious clinical findings in CKD can be explained by the structural and physiologic brain abnormalities of increased elevated cerebral blood flow (CBF), altered cerebral neurochemical concentrations and decreased white matter integrity.^3-11 12-15^

Patients with CKD have elevated cerebral blood flow (CBF),^7, 8^ likely secondary to disrupted cerebral autoregulation from dysfunctional blood brain barrier due to endothelial inflammation.^16, 17^ In addition, hemodialysis causes acute changes in the CBF, associated with ultrafiltration volume and change in hematocrit – major determinants of blood viscosity.^18^ These changes in cerebral hemodynamics can contribute to increase in stroke.^5, 19, 20^ Patients with CKD also have dysregulation of cerebral neurochemicals. Cerebral osmolytes such as myoinositol (ml) and choline containing compounds (Cho) are elevated in CKD.^4, 9-11^ Although causality is unproven, lower estimated glomerular filtration rate (eGFR) is correlated with higher concentrations of some of these osmolytes.^21^ Increase in cerebral osmolytes can alter cerebral osmotic pressure and impair cellular structure and function.^22^ In addition, patients with CKD have decreased white matter integrity measured by diffusion tensor imaging (DTI),^12-15^ a magnetic resonance imaging (MRI) method to measure the diffusion of water molecules along a nerve tract.^23^

Each of these abnormalities is associated with cognitive impairment in the non-CKD population,^24, 25^ and may be responsible for the high prevalence of cognitive impairment in ESKD.^26^ While many conditions associated with cognitive impairment, such as Alzheimer’s disease and traumatic brain injury have irreversible brain abnormalities,^27-30^ due to the unique nature of brain disease in CKD, it is possible that some of these abnormalities are reversible. Since cognition improves after KT,^31^ we hypothesize that brain abnormalities will also improve with KT. We used KT to comprehensively characterize the structural and physiological brain abnormalities in ESKD and determine their reversibility with KT. We examined CBF, cerebral neurochemical concentrations and white matter integrity longitudinally in patients before and after KT using non-invasive modern MRI techniques. This study builds upon the results of our prior study^31^ and reports a comprehensive evaluation of brain abnormalities pre-to post-KT.

## Methods

This is a single center prospective longitudinal observational cohort study on the effect of KT on brain health of ESKD patients on the KT waitlist in comparison to healthy non-CKD controls. The study was approved by the institutional review board and is registered in the US National Library of Medicine, www.clinicaltrials.gov (NCT01883349). Patients waitlisted for KT were enrolled and followed for 12 months post-KT. Brain MRI was performed at baseline, three months and 12 months post-KT. If patients did not receive KT within one year of the baseline visit, a second pre-KT brain MRI was performed to keep the time period between the last pre-KT MRI and KT to less than one year. Controls underwent brain MRI once.

### Participants

Adult participants between the ages of 30 and 70 who were listed for KT and expected to receive a KT within a year were enrolled. These included patients scheduled for a living donor KT and patients waitlisted for at least two years. Exclusion criteria included multi-organ listing, claustrophobia, MRI contraindications, recent stroke, uncontrolled psychosis, active seizure disorder, or current use of antipsychotics or anti-epileptics. Controls were age matched healthy persons without CKD or stroke and without contraindications to MRI. All participants signed informed consent prior to initiating study procedures.

### Demographics and clinical data

Demographic and clinical data were obtained from patients’ medical records and interviews. Demographic data included age, race, sex and education. Clinical data included assessment of comorbidities, specifically history of coronary artery disease (defined as history of myocardial infarction, coronary angioplasty or coronary artery bypass grafting), diabetes (defined as past or current use of oral hypoglycemics or insulin), hypertension (defined as past or current use of antihypertensives), stroke, depression, smoking (in the last 100 days), use of anticoagulants, atrial fibrillation, primary cause of ESKD and dialysis modality. The type of KT, time on dialysis before KT, kidney function at the time of KT for pre-emptive KTs, induction, panel reactive antibody, delayed graft function and episodes of rejection were also recorded for patients who received a KT during the study period.

Blood pressure, heart rate, and body mass index were also measured. At each visit, laboratory results including hemoglobin, serum creatinine, and tacrolimus levels were recorded from the latest values within the last three months. Serum hemoglobin and creatinine for controls were obtained from their primary care provider or tested specifically for the study to avoid enrollment of controls with undiagnosed CKD. Controls with estimated glomerular filtration rate of less than 60ml/min/1.73m^2^ were excluded from the study.

### MRI data acquisition

All MRI scans were performed using a 3 Tesla whole body scanner with a 20-channel head/neck receiver coil (Siemens Skyra, Erlangen, Germany). Modalities captured are described below. Acquisition parameters for each modality were designed to collect data from the whole brain, while maintaining good data quality, high signal to noise ratio, and acceptable spatial resolution.

#### CBF

CBF was measured using a pulsed arterial spin labeling (ASL) sequence optimized to measure blood flow in gray matter (repetition time/echo time (TR/TE) = 2500/12 ms, field of view (FOV) = 256 × 256 mm^2^, flip angle = 90 deg, matrix = 64 × 64, slice thickness/gap = 8.0/2.0 mm, voxel in-plane resolution = 4 × 4 mm^2^, 9 axial slices) acquiring 115 pairs of alternating labeled and unlabeled images.^32, 33^ The label offset was set to the distance between magnet isocenter and the junction of the cavernous and cerebral portions of the internal carotid arteries. However, in a subset of sessions, images were acquired so that the superior slice was at the vertex of the brain. We accounted for this variance by including slice position (upper or lower) as a covariate in the statistical analysis.

#### Cerebral neurochemicals

Neurochemical concentrations were measured with magnetic resonance spectroscopic imaging (MRSI) using a multi-voxel point resolved spectroscopy (PRESS) localized sequence (TR/TE = 1500/30 ms, FOV = 160 × 160 mm^2^, area of interest = 80 × 80 mm^2^, matrix = 16 × 16, final matrix size after 2x zero padding = 32 × 32, elliptical sampling, slice thickness = 10 mm, voxel in-plane resolution = 10 × 10 mm^2^).^34, 35^ MRSI data were acquired from a fronto-parietal slab placed superior to the corpus callosum and parallel to the anterior commissure-posterior commissure plane, which was positioned to cover the largest two-dimensional area of brain possible. The MRSI slab was carefully placed based on anatomical features in the mid-sagittal T_1_-weighted anatomic image to ensure that data were collected from the same anatomic location across participants at repeated time points. B_0_ shimming was performed using a vendor-provided field mapping technique.

#### White matter integrity

White matter integrity was measured by diffusion tensor imaging (DTI).^36, 37^ Data from the whole brain were collected using a double-refocused spin echo sequence optimized to capture major white matter tracts (TR/TE = 10,000/90 ms, FOV 300 × 300 mm, flip angle = 90 deg, matrix = 128 × 128, slice thickness = 2 mm, voxel in-plane resolution = 2.34 × 2.34 mm, 75 axial slices). The diffusion gradients were applied along 64 directions (diffusion gradient pulse δ = 14 ms, diffusion application time Δ = 53 ms, b value = 1,000 s/mm^2^).

#### T_1_ and T_2_*

T_1_- and T_2_*-weighted anatomic images were also collected and used to align the CBF, MRSI, and DTI data, and for normalization of the regional atlas masks. T_1_-weighted images were acquired with a 3D magnetization-prepared rapid acquisition gradient echo (MPRAGE) sequence (TR/TE = 2300/2.98 ms, FOV = 240 × 256 mm, flip angle = 9 deg, matrix = 240 × 256, slice thickness = 1.2 mm, voxel in-plane resolution = 1.0 × 1.0 mm, 176 sagittal slices).^38^ T_2_*-weighted images were utilized as the primary anatomic dataset for DTI alignment procedures. These images were acquired with a 2D fast low-angle shot (FLASH) sequence (TR/TE = 650/20 ms, FOV = 220 × 220 mm, flip angle = 20 deg, matrix = 256 × 256, slice thickness = 4.0 mm, voxel in-plane resolution = 0.86 × 0.86 mm, 44 axial slices).^39^

### MR Data Analysis

#### CBF

ASL data were processed using the ASLtbx^40^ with SPM12,^41^ modified for pulsed ASL. For each session, labeled and unlabeled ASL images were independently motion corrected and then a combined mean image was computed. The mean image was co-registered to match the T_1_-weighted anatomical image. The ASL images were then temporally filtered and spatially smoothed with a 6 mm full-width half-maximum Gaussian kernel. CBF was then estimated by subtraction of labeled and unlabeled mean images, resulting in a mean CBF image (units = ml/min/100mg tissue). The structural correlation based outlier rejection (SCORE) algorithm was used to remove outlier voxels.^42^ The T_1_-weighted scan was then normalized using unified segmentation-normalization (SPM12). In addition to whole gray matter CBF, we also assessed regional CBF. These regions were selected *a priori* based on atlas-based regional parcellations of the whole gray matter area distributed across the entire brain. For regional CBF, we used 14 gray matter regions from the Automated Anatomical Labeling atlas; anterior cingulate cortex, middle frontal gyrus, hippocampus, primary motor cortex, posterior cingulate cortex, precuneus, superior parietal cortex, temporal cortex, thalamus, pallidum, putamen, caudate, frontal cortex, and parietal cortex.^43^ Mean CBF was computed in regions of interest created from the intersection of the individual gray matter segmentation (probability > 0.5) performed under the SPM unified segmentation algorithm and regional masks from the Automated Anatomical Labeling atlas. Since calculation of CBF includes the MRI longitudinal relaxation time (T1) of blood as a variable, and T1 of blood varies with hematocrit, the calculation was corrected for hematocrit.^44^

#### Neurochemicals

MRSI data were analyzed using LCModel^45^ with a numerically simulated basis set for concentrations of N-acetylaspartate (NAA), Cho, glutamate and glutamine (Glx), mI and total creatine (Cr). Metabolite concentrations were extracted from gray matter within the MRSI acquisition slab. Voxels were selected for analysis based on the tissue fraction (gray/white matter) in each voxel using the segmented T_1_-weighted images and slice selection profiles of radiofrequency pulses corresponding to the spectroscopic voxels. Voxels were included in the analysis if the tissue fraction was greater than 50% gray matter. Analysis was further limited to the center of the region to capture the best quality data and reduce off-resonance effects from the skull and fat surrounding the head. Final concentrations of neurochemicals were therefore obtained from the central 50 × 50 mm^2^ region of the acquisition slab by averaging concentration values using weighting factors derived from the goodness of LCModel fit.^46^ To correct for the differences in levels of different neurochemicals, we normalized the values within each individual by reporting concentrations for NAA, Cho, mI and Glx as ratios to Cr.^47^

#### White matter integrity

DTI data were processed using Analysis of Functional NeuroImages (AFNI)^48^ software, and TORTOISE version 3.1.4 DIFF_PREP and DIFF_CALC processing modules. Processing steps included eddy current correction modeled with quadratic functions; motion distortion correction; echo planar imaging distortion correction; Gibbs ringing correction;^49^ and denoising. TORTOISE DIFF_CALC EstimateTensorNLLS was used to fit the tensors. DIFFCALC GUI version 2.5 running under IDL Virtual Machine was used to compute the eigenvectors, derive the tensors, and save the output metrics.

DTI output metrics included fractional anisotropy (FA), a measurement of whether diffusion is constrained along any axis, indicating the presence of white matter fiber tracts (range 0-1, with 0 indicating equal diffusion in all directions and 1 indicating unidirectional diffusion), and mean diffusivity (MD), which is calculated by dividing the total diffusion for all three eigenvectors at each voxel by three (for the three eigenvectors, units = 10^−3^ mm^2^/s^−1^). Diffusivity is reduced in constrained areas like dense white matter fiber tract bundles. High MD may indicate more free water or edema in the brain.^23^ Mean FA and MD were exported for each participant and session from regional white matter tract masks. We used 11 white matter tracts from the Johns Hopkins University DTI-based white matter atlases; anterior thalamic radiation, cingulum in the cingulated cortex areas, cingulum in the hippocampal area, corticospinal tract, forceps major, forceps minor, inferior fronto-occipital fasciculus, inferior longitudinal fasciculus, superior longitudinal fasciculus, temporal projection of the superior longitudinal fasciculus, and uncinate fasciculus.^50-52^ A combined All-tract mask was created as the overlap of all regional masks.

##### Primary and secondary outcomes

Our primary outcomes were the effects of KT on whole brain gray matter CBF, cerebral neurochemicals (NAA/Cr, Cho/Cr, Glx/Cr, and mI/Cr), and DTI-measured whole brain white matter FA and MD. As secondary outcomes, to better understand possible anatomic specificity of changes, we measured regional CBF, FA, and MD in atlas-defined anatomic regions distributed across the entire brain.

## Statistical Analysis

Baseline characteristics between patients and controls were analyzed using descriptive statistics. For categorical variables, differences in frequencies were measured using a nonparametric Fisher exact test. For continuous variables, mean differences were measured using a two sample t-test. Assumptions of normality for the t-test were inspected by the quantile-quantile plot and histogram. For clinical measurements, unadjusted one-way repeated analysis of variance (ANOVA) was used to measure the continuous outcomes between pre- and post-KT patient groups. Residuals were assessed to evaluate the fit of the underlying model assumptions.

Each imaging modality (ASL, MRSI and DTI) was analyzed separately. Imaging data in controls, as well as pre- and post-KT groups were adjusted for covariates of age,^53^ race,^54^ sex,^55^ and education level^56, 57^ that can confound results. Since the exact time for KT is unpredictable, a linear mixed-model approach was used to account for repeated observations on participants. A linear mixed-model is more flexible than repeated-measures ANOVA; it allows for variability in time between scans, variability in the number of scans per participant, and the inclusion of participants who have missing data at one or more timepoints.^58, 59^ Each participant’s individual intercept was estimated in the model using the random intercept term. Age was calculated at every timepoint in order to both control for the effect of age and function as the time variable. All available imaging data were included in the model to detect any group differences between pre-KT, post-KT, and controls. The pre-KT group includes both pre-KT scans, if available; the post-KT group includes both the 3 month and 12 month post-KT scans, if available. The effect of group (control, pre-KT, post-KT) and covariates (age, race, sex, and education level) were included as fixed effects. Residual plots were assessed to evaluate the model fit. We performed pairwise testing of the three groups, using F- and t-tests as appropriate. We constructed scatter plots mapping each participant’s observed measures over time (chronological age), showing both the overall change with aging and the effect of KT. Kidney function (eGFR and serum creatinine) was correlated with post-KT CBF using Pearson’s correlation. As a test for whether there would be any change associated with time alone, for those patients who had repeated pre-KT MRI, the pre-KT scans were compared using Wilcoxon nonparametric signed rank test. Results with p<0.05 were considered statistically significant. All statistical analyses were performed using R Studio (Version 3.6.3) and SAS (Version 9.4, The SAS institute, Cary, NC).

## Results

### Demographic and Clinical Comparisons

A total of 48 participants, 29 patients and 19 controls were enrolled (**Table 1**). Of the 29 patients, 23 received a KT. 22 completed the 3 month post-KT MRI and 18 completed the 12 month post-KT MRI. Four of the 29 patients were not transplanted within one year and per study protocol, underwent repeated baseline MRI at one year after enrollment. Patients and controls had no differences in age, race, sex or education (all p>0.10) (**Table 1**). The mean serum creatinine and eGFR for controls were 0.85±0.13 mg/dl and 92.8±13.5 ml/min respectively. ESKD patients had a greater incidence of diabetes (p=0.03) and hypertension (p<0.0001) compared to controls. The most common causes of ESKD were diabetes (21%) and autosomal dominant polycystic kidney disease (28%). Six patients had not initiated dialysis at baseline. Clinical characteristics pre- and post-KT are summarized in **Table 2**. Hematocrit and serum calcium increased and serum bicarbonate decreased after KT. Other parameters such as blood pressure, heart rate, and body mass index did not change with KT (all p>0.10) (**Table 2**).

**Table 1:** Baseline demographics and clinical characteristics of study participants.

|  | Controls<br>(n=19) | ESKD patients<br>(n=29) | p-value |
| --- | --- | --- | --- |
| **Age years, mean ± SD** | 48.78 ± 8.38 | 53.43 ± 11.38 | 0.11 |
| **Race, n (%)** |  |  | 0.65 |
| White | 17 (89.5) | 25 (86.2) |  |
| Black | 0 (0.0) | 2 (6.9) |  |
| Other | 2 (10.5) | 2 (6.9) |  |
| **Sex n (%)** |  |  | 0.23 |
| Males | 9 (47.4) | 20 (69.0) |  |
| Females | 10 (52.6) | 9 (31.0) |  |
| **Education, n (%)** |  |  | 0.32 |
| High school diploma | 1 (5.3) | 5 (17.2) |  |
| Some college | 3 (15.8) | 9 (31.0) |  |
| 4-year college degree | 7 (36.8) | 8 (27.6) |  |
| Graduate school | 8 (42.1) | 7 (24.1) |  |
| **Comorbid conditions, n (%)** |  |  |  |
| Coronary artery disease | 0 (0.0) | 3 (10.3) | 0.27 |
| Diabetes | 0 (0.0) | 7 (24.1) | 0.03 |
| Hypertension | 5 (26.3) | 25 (86.2) | <0.0001 |
| Stroke | 0 (0.0) | 1 (3.5) | 0.99 |
| Depression | 2 (10.5) | 7 (24.1) | 0.29 |
| Smoking | 6 (31.6) | 9 (31.0) | 0.99 |
| **On anticoagulants, n (%)** |  | 2 (6.9) |  |
| **Atrial fibrillation, n (%)** |  | 3 (10.3) |  |
| **Cause of ESKD, n (%)** | NA |  | NA |
| Diabetes |  | 6 (20.7) |  |
| Hypertension |  | 3 (10.3) |  |
| ADPKD |  | 8 (27.6) |  |
| Other |  | 12 (41.4) |  |
| **Dialysis modality, n (%)** | NA |  | NA |
| In center hemodialysis |  | 11 (37.9) |  |
| Home hemodialysis |  | 3 (10.4) |  |
| Peritoneal dialysis |  | 9 (31.0) |  |
| Not on dialysis |  | 6 (20.7) |  |

ESKD: end-stage kidney disease, ADPKD: autosomal dominant polycystic kidney disease. Continuous values are presented as mean ± standard deviation (SD), p values represent unadjusted two sample t-test. Categorical variables are presented as frequency (percentage), p value represents unadjusted nonparametric Fisher exact test.

**Table 2:** Pre- to post-KT changes in clinical measures.

|  | **1-year pre-KT**<br>**(n=4)**<br>**(mean ± SD)** | **Pre-KT**<br>**(n=29)**<br>**(mean ± SD)** | **3 months post-KT**<br>**(n=22)**<br>**(mean ± SD)** | **12 months post-KT**<br>**(n=18)**<br>**(mean ± SD)** | **p-value** |
| --- | --- | --- | --- | --- | --- |
| **Hematocrit** | 33.9 ± 4.6 | 33.3 ± 4.1 | 39.7 ± 4.2 | 43.6 ± 4.2 | <0.0001 |
| **Serum calcium (mg/dl)** | 9.5 ± 0.4 | 9.5 ± 0.5 | 9.8 ± 0.5 | 9.7 ± 0.4 | 0.004 |
| **Serum bicarbonate (mEq/L)** | 23.3 ± 2.2 | 25.5 ± 3.5 | 24.0 ± 1.8 | 23.3 ± 2.7 | 0.02 |
| **Serum Creatinine (mg/dl)** | NA | NA | 1.4 ± 0.5 | 1.3 ± 0.5 | NA |
| **eGFR (mL/min/1.73m<sup>2</sup>)** | NA | NA | 55.4 ± 13.7 | 55.7 ± 13.8 | NA |
| **Tacrolimus level (ng/mL)** | NA | NA | 11.4 ± 12.4 | 7.8 ± 3.2 | NA |
| **SBP (mm Hg)** | 114.5 ± 7.0 | 141.9 ± 16.5 | 137.5 ± 17.8 | 138.8 ± 15.1 | 0.69 |
| **DBP (mm Hg)** | 62.3 ± 10.5 | 79.2 ± 10.3 | 77.7 ± 10.1 | 77.3 ± 13.2 | 0.98 |
| **HR (beats/min)** | 75.5 ± 7.7 | 75.9 ± 14.5 | 75.4 ± 10.8 | 74.4 ± 16.1 | 0.67 |
| **BMI (kg/m<sup>2</sup>)** | 31.0 ± 3.8 | 30.0 ± 4.8 | 30.0 ± 5.1 | 35.0 ± 13.2 | 0.20 |

KT: kidney transplantation, eGFR: estimated glomerular filtration rate, SBP: systolic blood pressure, DBP: diastolic blood pressure, HR: heart rate, BMI: body mass index.
All measures are presented as mean ± standard deviation (SD).
p values represent unadjusted one-way repeated measures ANOVA for pre-to post-transplant comparison.

All transplanted patients received induction and maintenance immunosuppression per institutional protocol. **Supplementary Table 1** describes the clinical characteristics of the 22 patients who had a post-KT MRI. Per institutional policy, ABO-incompatible KTs were not performed at our center. Induction immunosuppression consisted of either thymoglobulin or basiliximab (depending on immunological risk of the patient), steroids and a mycophenolate compound. A calcineurin inhibitor (tacrolimus) was started 24 hours after the surgery. Maintenance immunosuppression consisted of a mycophenolate compound and tacrolimus with or without low dose steroids. Post-KT, all patients were on tacrolimus and had a functional graft with a mean serum creatinine of 1.4±0.5 mg/dl (**Table 2**).

### Cerebral blood flow

**Table 3** shows the comparisons of adjusted ASL-measured CBF between different groups over the total gray matter and in specific anatomic regions within the total gray matter mask. CBF in the total gray matter was higher in pre-KT ESKD patients compared to controls (p=0.003) **(Table 3 and Figure 1)** and normalized post-KT to values observed in controls. **Figure 1** shows data for individual participants at pre- and post-KT for comparison. Normalization of CBF post-KT did not correlate with eGFR (r=0.16, p=0.47) or serum creatinine (r=-0.29, p=0.19) **(Supplementary Figure 1)**. When CBF was analyzed regionally, the decrease in CBF with KT was consistent across all brain regions **(Table 3)**.

**Table 3:** Overall gray matter and regional comparisons of cerebral blood flow in pre-KT ESKD patients, post-KT ESKD patients and controls.

|  | p-value <sup>a</sup> | Pairwise comparisons<br>Estimated difference (p-value) <sup>b</sup> |  |  |
| --- | --- | --- | --- | --- |
|  |  | Change pre- to<br>post-KT | Pre-KT vs. control | Post-KT vs. control |
| Total gray matter | <0.0001 | -7.24 (<0.0001) | 7.91 (0.003) | 0.67 (0.78) |
| **Regions analyzed** |  |  |  |  |
| ACC | 0.07 | -5.40 (0.02) | 5.21 (0.20) | -0.19 (0.96) |
| Frontal Mid | 0.008 | -7.37 (0.002) | 7.00 (0.05) | -0.37 (0.91) |
| Hippocampus | <0.0001 | -8.67 (<0.0001) | 10.89 (0.002) | 2.21 (0.46) |
| M1 | 0.002 | -6.30 (0.001) | 7.58 (0.02) | 1.28 (0.65) |
| PCC | 0.0004 | -12.17 (<0.0001) | 9.39 (0.06) | -2.78 (0.54) |
| Precuneus | 0.0001 | -10.40 (<0.0001) | 9.12 (0.017) | -1.28 (0.71) |
| Superior Parietal | 0.03 | -6.77 (0.01) | 4.45 (0.24) | -2.32 (0.49) |
| Temporal | 0.001 | -5.84 (0.001) | 7.69 (0.01) | 1.85 (0.47) |
| Thalamus | <0.0001 | -2.55 (<0.0001) | 13.15 (0.01) | 0.28 (0.95) |
| Pallidum | 0.03 | -6.11 (0.01) | 5.77 (0.17) | -0.35 (0.93) |
| Putamen | 0.001 | -7.09 (0.001) | 8.26 (0.018) | 1.17 (0.71) |
| Caudate | 0.008 | -7.13 (0.002) | 6.63 (0.05) | -0.49 (0.87) |
| Frontal | 0.003 | -6.30 (0.001) | 7.26 (0.01) | 0.96 (0.71) |
| Parietal | 0.0003 | -8.36 (<0.0001) | 8.85 (0.01) | 0.49 (0.87) |
| KT: kidney transplantation, ACC: anterior cingulate cortex, Frontal Mid: middle frontal gyrus, M1: primary motor cortex, PCC: posterior cingulate cortex. |  |  |  |  |
| <sup>a</sup> p value for linear mixed model F-test for any group differences between pre-KT, post-KT, and controls, adjusted for age, race, sex, and education level. Pre-KT includes both pre-KT scans, if available; post-KT includes both the 3 month and 12 month post-KT scans, if available. <sup>b</sup> p value for adjusted linear contrast of pairwise estimated group differences t-test. All units are in ml/min/100mg tissue. |  |  |  |  |

**Figure 1:**
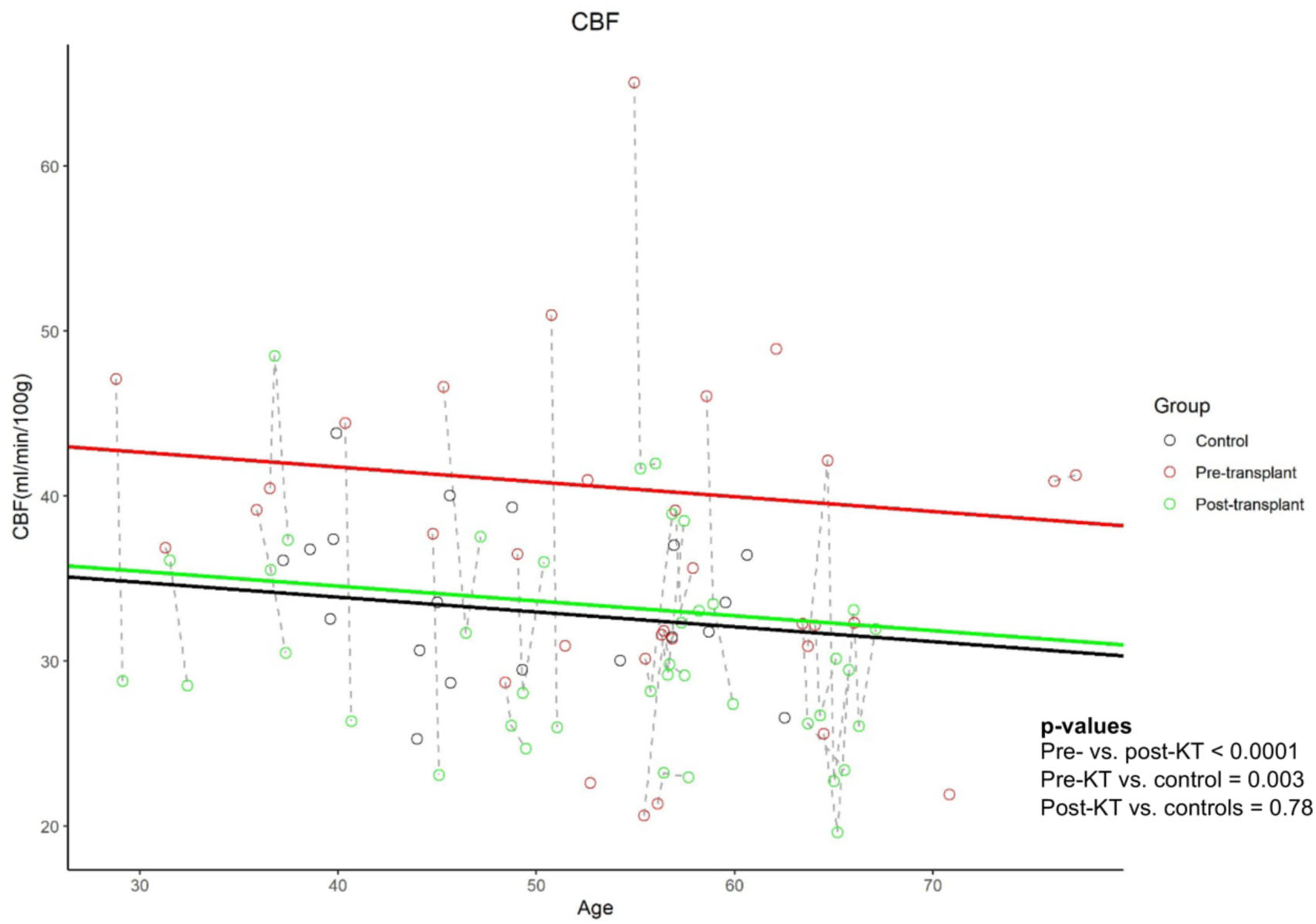
Comparisons of cerebral blood flow in total gray matter in pre-KT patients, post-KT patients and controls. Scatterplot displays individual participant data as a function of age and group (Controls: Black circles, Pre-KT: Red circles, Post-KT, Green circles). Dashed lines represent individual participant trajectory over time. Distance between the solid lines represents the estimated group mean differences for fixed covariate values of race (white), sex (male), education (more than high school), and slice position (upper). The slope of the solid lines represents the overall effect of age. ESKD: end stage kidney disease, KT: kidney transplantation, CBF: cerebral blood flow.

### Brain neurochemicals

**Table 4** shows the group comparisons of different neurochemicals measured with MRSI in the in our adjusted linear mixed model. **Figure 2** shows data for individual patients at different time points in the study. Of the four neurochemicals that were analyzed, the Cho/Cr (p=0.001) and ml/Cr (p=0.0001) were higher in pre-KT patients compared to controls and normalized post-KT. NAA/Cr and Glx/Cr were not different between the pre-KT patients and controls and did not change post-KT.

**Table 4:** Comparisons of brain neurochemical concentrations normalized to creatine in pre-KT ESKD patients, post-KT ESKD patients and controls.

| Neurochemical analyzed | p-value <sup>a</sup> | Pairwise comparisons<br>Estimated difference (p-value) <sup>b</sup> |  |  |
| --- | --- | --- | --- | --- |
|  |  | Change pre- to post-KT | Pre-KT vs. control | Post-KT vs. control |
| **NAA/Cr** | 0.04 | 0.02 (0.14) | 0.06 (0.08) | 0.09 (0.02) |
| **Cho/Cr** | <0.0001 | -0.006 (<0.0001) | 0.04 (0.001) | 0.008 (0.47) |
| **Glx/Cr** | 0.23 | 0.05 (0.23) | -0.08 (0.10) | -0.04 (0.47) |
| **ml/Cr** | <0.0001 | -0.06 (0.0003) | 0.11 (0.0001) | 0.05 (0.05) |

KT: kidney transplantation, NAA: N-acetylaspartate, Cr: total creatine, Cho: choline, Glx: glutamate and glutamine, ml: myo-inositol.
<sup>a</sup> p value for linear mixed model F-test for any group differences between pre-KT, post-KT, and controls, adjusted for age, race, sex, and education level. Pre-KT includes both pre-KT scans, if available; post-KT includes both the 3 month and 12 month post-KT scans, if available. <sup>b</sup> p value for adjusted linear contrast of pairwise estimated group differences t-test. Units are ratios to creatine.

**Figure 2:**
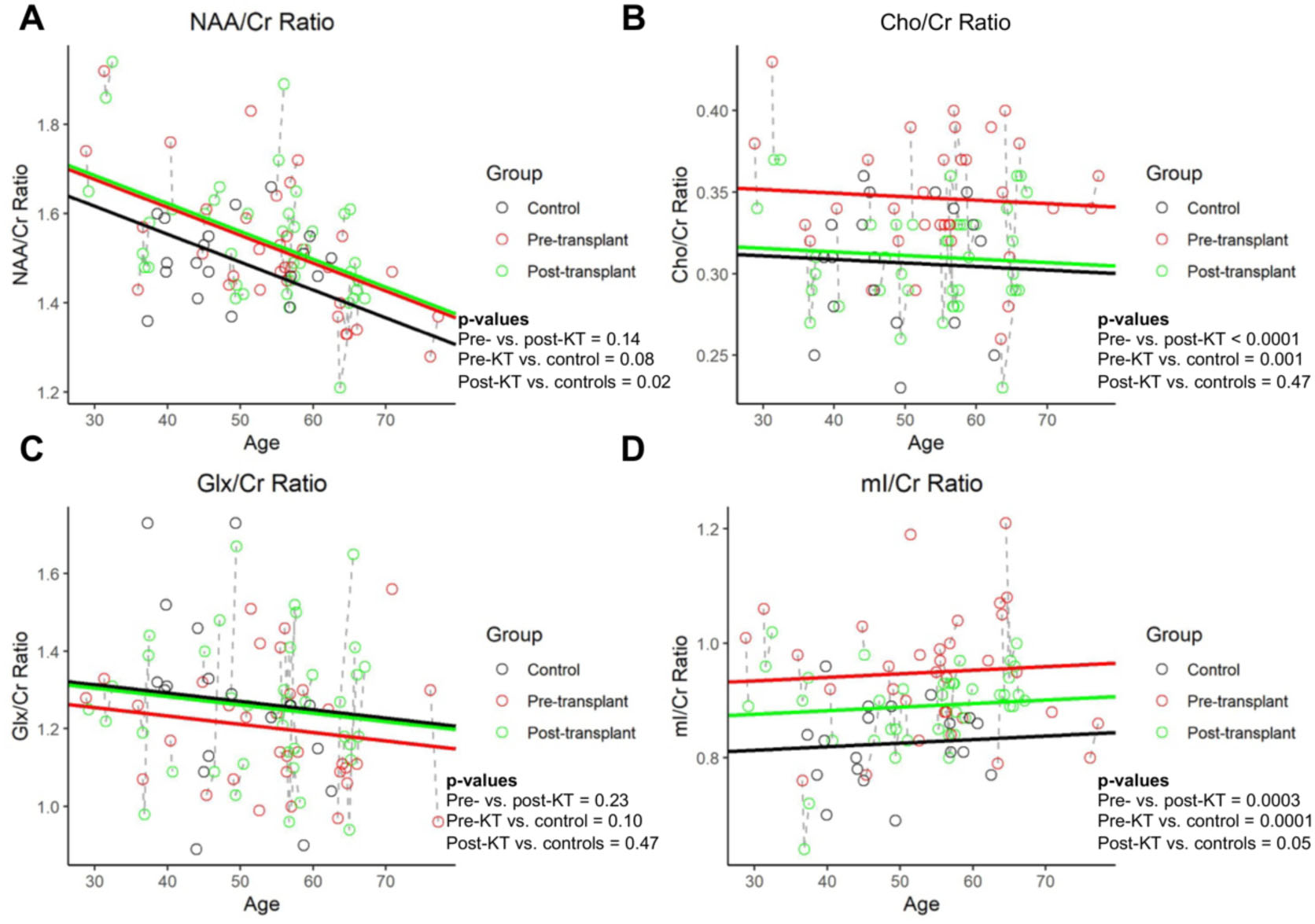
Comparisons of MR spectroscopy imaging measured **A)** NAA/Cr ratio **B)** Cho/Cr ratio **C)** Glx/Cr ratio and **D)** ml/Cr ratio in pre-KT patients, post-KT patients and controls. Scatterplot displays individual participant data as a function of age and group (Controls: Black circles, Pre-KT: Red circles, Post-KT, Green circles). Dashed lines represent individual participant trajectory over time. Distance between the solid lines represents the estimated group mean differences for fixed covariate values of race (white), sex (male), and education (more than high school). The slope of the solid lines represents the overall effect of age. ESKD: end stage kidney disease,

### White matter integrity

**Table 5 and Figure 3** show adjusted group comparisons of FA and MD for all tracts and for each of the regional tracts. FA increased pre- to post-KT (p=0.001). Neither pre- (p=0.23) nor post-KT (p=0.72) FA was different from controls. MD decreased post-KT (p=0.0001). Similar to FA, neither pre-KT (p=0.33) nor post-KT (p=0.89) MD was different from controls.

**Table 5:** Whole brain and regional comparisons of diffusion metrics fractional anisotropy (FA) and mean diffusivity (MD) in pre-KT ESKD patients, post-KT ESKD patients and controls.

| Regions and metrics analyzed | p-value <sup>a</sup> | Pairwise comparisons<br>Estimated difference (p-value) <sup>b</sup> |  |  |
| --- | --- | --- | --- | --- |
|  |  | Change pre- to post-KT | Pre-KT vs. control | Post-KT vs. control |
| All tracts FA | 0.003 | 0.007 (0.001) | -0.010 (0.23) | -0.003 (0.72) |
| All tracts MD | 0.001 | -0.022 (0.0001) | 0.019 (0.33) | -0.003 (0.89) |
| **Regional FA** |  |  |  |  |
| ATR | 0.001 | 0.011 (0.0002) | -0.018 (0.19) | -0.007 (0.63) |
| CG | 0.71 | 0.004 (0.58) | -0.024 (0.52) | -0.020 (0.59) |
| CH | 0.51 | 0.109 (0.46) | 0.017 (0.52) | 0.028 (0.29) |
| CST | 0.06 | 0.009 (0.02) | -0.001 (0.93) | 0.008 (0.50) |
| FMAJ | 0.26 | 0.006 (0.14) | -0.015 (0.40) | -0.009 (0.62) |
| FMIN | 0.22 | 0.006 (0.16) | 0.008 (0.45) | 0.014 (0.19) |
| IFOF | 0.02 | 0.008 (0.01) | -0.015 (0.20) | -0.007 (0.56) |
| ILF | 0.005 | 0.009 (0.002) | -0.015 (0.16) | -0.006 (0.58) |
| SLF | 0.23 | 0.002 (0.54) | -0.015 (0.09) | -0.013 (0.14) |
| SLFT | 0.60 | 0.002 (0.52) | -0.010 (0.40) | -0.007 (0.52) |
| UF | 0.27 | 0.011 (0.15) | -0.017 (0.31) | -0.007 (0.70) |
| **Regional MD** |  |  |  |  |
| ATR | 0.008 | -0.367 (0.002) | 0.045 (0.47) | 0.008 (0.90) |
| CG | 0.0018 | -0.016 (0.005) | 0.020 (0.25) | 0.004 (0.83) |
| CH | 0.33 | -0.169 (0.50) | -0.070 (0.25) | -0.087 (0.15) |
| CST | 0.12 | -0.011 (0.09) | -0.013 (0.41) | -0.024 (0.14) |
| FMAJ | 0.004 | -0.042 (0.002) | 0.049 (0.15) | 0.007 (0.83) |
| FMIN | 0.08 | -0.020 (0.03) | 0.002 (0.92) | -0.018 (0.35) |
| IFOF | 0.0003 | -0.187 (0.0001) | 0.027 (0.11) | 0.009 (0.60) |
| ILF | 0.003 | -0.006 (0.001) | 0.026 (0.35) | 0.002 (0.93) |
| SLF | 0.003 | -0.012 (0.001) | 0.022 (0.13) | 0.009 (0.52) |
| SLFT | 0.0003 | -0.010 (0.0001) | 0.015 (0.12) | 0.005 (0.59) |
| UF | 0.33 | -0.020 (0.14) | 0.009 (0.82) | -0.011 (0.78) |

KT: kidney transplantation, FA: fractional anisotropy, MD: mean diffusivity, ATR: anterior thalamic radiation, CG: cingulum in the cingulate cortex area CH: cingulum in the hippocampal area, CST: corticospinal tract, FMAJ: forceps major, FMIN: forceps minor, IFOF: inferior fronto-occipital fasciculus, ILF: inferior longitudinal fasciculus, SLF: superior longitudinal fasciculus, SLFT: temporal projection of the SLF, UF: uncinate fasciculus.
<sup>a</sup> p value for linear mixed model F-test for any group differences between pre-KT, post-KT, and controls, adjusted for age, race, sex, and education level. Pre-KT includes both pre-KT scans, if available; post-KT includes both the 3 month and 12 month post-KT scans, if available. <sup>b</sup> p value for adjusted linear contrast of pairwise estimated between group differences t-test. Units are scalar (0-1) for FA and $10^{-3} \text{ mm}^2/\text{sec}^{-1}$ for MD.

**Figure 3:**
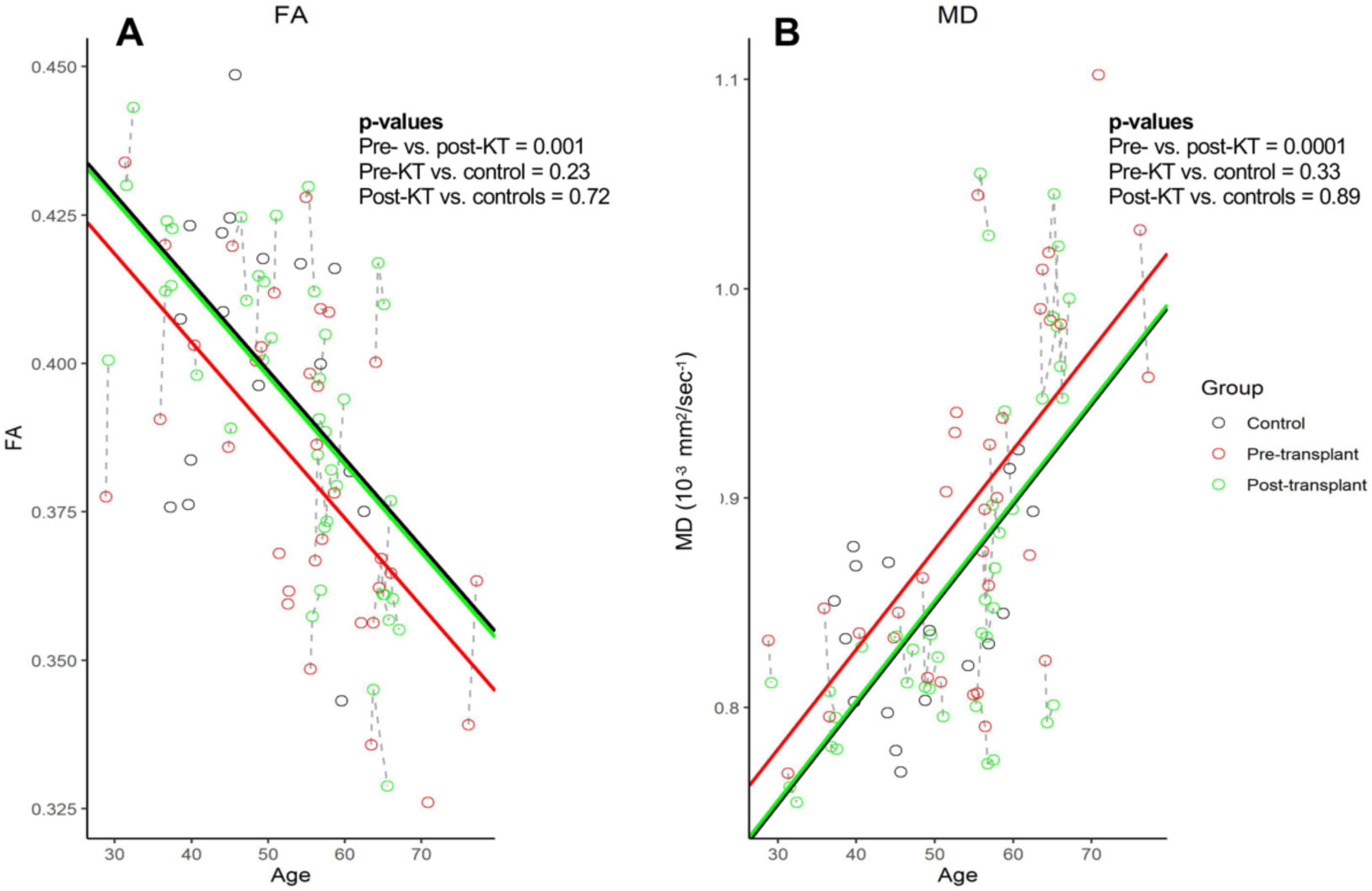
Comparisons of diffusion metrics **A)** Fractional anisotropy (FA) and **B)** Mean diffusivity (MD) in whole brain in pre-KT patients, post-KT patients and controls. Scatterplot displays individual participant data as a function of age and group (Controls: Black circles, Pre-KT: Red circles, Post-KT, Green circles). Dashed lines represent individual participant trajectory over time. Distance between the solid lines represents the estimated group mean differences for fixed covariate values of race (white), sex (male), and education (more than high school). The slope of the solid lines represents the overall effect of age. ESKD: end stage kidney disease, KT: kidney transplantation, FA: fractional anisotropy, MD: mean diffusivity.

### Brain changes without KT

Out of the 29 ESKD patients in the study, four patients had a second pre-KT MRI while awaiting KT. There was no change in CBF, cerebral neurochemicals, FA or MD over one year in these patients (all p>0.10) **(Supplementary Table 2)**.

## Discussion

In this study we found that structural and physiologic brain abnormalities in ESKD reversed post-KT. CBF, which was elevated pre-KT, decreased post-KT to levels in controls, globally as well as in all anatomic regions of gray matter analyzed. Cho/Cr and ml/Cr which were also elevated pre-KT normalized post-KT. Finally, white matter integrity also normalized with increase in FA and decrease in MD post-KT. We also noted the expected age-related changes in brain measurements that were controlled for in our linear mixed model.

The reversibility of brain abnormalities in CKD has important mechanistic and therapeutic implications. Potential reversibility underscores the need to develop improved management strategies other than KT for patients who cannot be transplanted. Better dialysis techniques that include targeting dialysis related ischemia to reduce metabolic derangements could help prevent and perhaps mitigate some of these abnormalities. Preservation of residual renal function to retain tubular secretion may also positively impact these brain abnormalities. The overarching impact of our observations would be to identify patients with reversible vs. irreversible brain abnormalities and to devise strategies to reverse brain abnormalities with the goal of improving cognition. Cognitive impairment affects eligibility for KT.^60^ However, given that cognitive impairment and brain abnormalities will improve with KT, more of these patients who are currently rejected for KT may be able to receive a KT.

Elevated CBF seen in our pre-KT patients is consistent with other studies in adult^8, 17^ and pediatric^61^ patients with CKD. Disruption of cerebral autoregulation due to inflammation and endothelial dysfunction affecting the blood brain barrier may play a role.^16, 17^ Alternatively, elevated CBF could be secondary to increased metabolic demand. Our results that CBF decreased pre- to post-KT is consistent with prior cross-sectional^62^ and longitudinal studies.^63^ Some other studies found CBF to be decreased in CKD^64, 65^ likely due to differences in the methodology of assessing CBF. ASL has the advantage of repeatability and ability to quantify global and regional CBF.^66^ Since hemodialysis preferentially affects the watershed areas of the brain,^67^ we also explored regional changes in the brain in addition to global changes. All regions showed a decreasing trend in CBF **(Table 3)**, pointing to systemic etiologies such as reduced inflammation and improvment of cerebral autoregulation. It is also possible that CBF decreased due to the vasoconstrictive effects of calcineurin inhibitors. Distinct from the KT-related change in CBF, we observed an age-related decrease in CBF in all participants, a well-established phenomenon seen in the general population.^68^ Although CBF correlates with eGFR in nontransplanted CKD,^8^ we did not see a strong correlation between CBF and post-KT serum creatinine or eGFR in our study **(Supplementary Figure 1)**. This is consistent with other studies in KT where CBF^62^ or cognition^69^ is independent of eGFR. It is possible that the changes in CBF and cognition are not caused directly by reduced eGFR, but by an additional confounding variable such as endothelial inflammation or microvascular disease that influences both eGFR and CBF.

Similar to other published studies, we found that pre-KT patients had higher cerebral Cho/Cr and ml/Cr compared to controls.^4, 9-11^ Both Cho/Cr and ml/Cr decreased post-KT. Cho is a phospholipid cell membrane precursor and represents breakdown products such as phosphocholine, phosphatidylcholine, glycerophosphorylcholine and phosphorylcholine.^22^ Importantly, Cho is also a precursor of trimethylamine N-oxide, a marker of cardiovascular disease and mortality.^70, 71^ ml, primarily found in glial cells, is involved in the phosphoinositide-mediated signal transduction and is a marker of inflammation or gliosis.^22^ High plasma ml is also associated with the high prevalence of peripheral polyneuropathy in ESKD.^72^ With their relative small size (molecular weight 105 g/mol and 180 g/mol respectively), both compounds are readily filtered by the glomeruli and eliminated with tubular secretion.^73, 74^ Both plasma^72, 75^ and intracerebral Cho and ml concentrations are elevated in CKD^21^ and decrease only slightly with dialysis.^9^ Cho and ml are both major cerebral osmolytes and high concentrations can increase osmotic pressure in the brain, cause cellular edema and alter cellular structure and function. Both compounds are transported from the plasma to the brain through the blood brain barrier by diffusion and carrier mediated transporter systems.^76, 77^ Due to the blood brain barrier, changes in plasma concentrations of neurochemicals do not correlate with cerebral concentrations in healthy individuals.^78^ However, with the disruption of the blood brain barrier in CKD, higher serum concentrations could theoretically affect cerebral neurochemical concentrations. Our study indicates normalization of Cho and ml post-KT, perhaps by better elimination with tubular secretion^73, 74^ which cannot be restored with dialysis. Improvement in these neurochemicals after KT could restore cerebral osmotic regulation, lower cellular edema and improve cell function and cognition.

Similar to a prior cross-sectional study,^9^ we did not see any differences in the levels of Glx/Cr and NAA/Cr between controls and pre-KT patients, suggesting that CKD does not impact these neurochemicals as much as Cho and ml. Consistent with this hypothesis, there was no change in Glx/Cr and NAA/Cr pre- to post-KT. Glutamine is mainly found in glia and is a precursor to glutamate,^22^ the most abundant excitatory neurotransmitter in the neurons. NAA is also an osmolyte located in the cell bodies, axons and dendrites of neurons and is a marker of neuronal number, density and integrity.^79^ NAA/Cr is often reduced in neurodegenerative diseases and may not be affected in CKD.^22^

Similar to our prior study,^31^ KT resulted in an increase in FA and a decrease in MD. A recent study demonstrated similar findings.^63^ These changes in FA and MD indicate decreased free dispersion and greater axial movement of water molecules along the white matter tracts. A decrease in cerebral osmolytes and improvement in cerebral edema as described above can explain these DTI results. Pre-KT, even with maintenance dialysis, the clearance of substances that are eliminated by tubular secretion is reduced and can result in (often subclinical) cerebral edema. Although we have previously shown that patients on dialysis have lower FA and higher MD compared to healthy controls,^80^ we did not find FA and MD values to be significantly different in pre-KT patients and controls in this study. This could be because of the small sample size and single MRI measurements in controls leading to a larger standard deviation.

The evidence that dialysis has short-term and long-term deleterious effects on brain health is rapidly accumulating.^4, 5 9-15, 19, 20^ Our MRSI and DTI results indicate normalization of cerebral edema post-KT, a finding that is not observed with dialysis. In fact, cerebral edema increases with longer dialysis vintage and higher ultrafiltration volumes.^81^ Similarly, change in CBF with dialysis can be detrimental.^2^ We demonstrate reversibility in abnormalities in CBF, neurochemical concentrations and white matter integrity in ESKD. Based on our study protocol, we repeated pre-KT MRI in patients who did not receive KT within one year. We were able to compare two pre-KT MRIs in these patients and did not observe any change in brain abnormalities. Although we do not have the power to make definite conclusions in this study, our previous work^80^ is consistent with this observation. Normalization of brain abnormalities even after 12 months post-KT indicates that the post-KT changes were not due to acute effects of KT such as steroids, but a phenomenon that persists beyond the immediate post-KT period.

Despite our modest sample size, our longitudinal approach, linear mixed model analysis, and a uniform change in measures among patients strengthen our findings. For example, all patients without exception had a decrease in CBF after KT. Unlike other studies, our CBF calculations were corrected for hematocrit. Hematocrit affects blood viscosity and the T1 of blood in MRI.^44^ As observed in this study, hematocrit changes following KT. Thus, correction for hematocrit is important for accurate CBF measurements in ESKD. Our models were adjusted for age,^53^ race,^54^ sex,^55^ and level of education.^56, 57^ Age in particular is associated with changes in brain volume and function, and age-related changes observed over many years could be larger than changes observed in 12 months post-KT. Another strength was the measurement of regional CBF, FA and MD changes in addition to global changes.

In summary, abnormalities in CBF, neurochemical concentrations and white matter integrity in CKD are normalized with KT. This reversibility in brain abnormalities has important implications in appreciating and managing the risk of dementia and stroke in our CKD population. More studies are needed to understand the mechanisms underlying these brain abnormalities, the role of residual kidney function in preserving brain health on dialysis, and to explore innovations in renal replacement therapies to mitigate these abnormalities even in patients who cannot be transplanted.

## Data Availability

Data are available by request from the corresponding author (A.G.).

## Author contributions

A.G., W.M.B., J.M.B. and M.J.S. designed the study. R.J.L., E.D.V., and I.Y.C. analyzed the MRI data. R.N.M., P.S. and J.D.M. did the statistical analysis. A.G. and R.J.L. drafted the manuscript. W.M.B., J.M.B., R.N.M., E.D.V. J.D.M. and M.J.S. reviewed and edited the manuscript. All authors approved the final version of the manuscript.

## Acknowledgments

This work was supported by National Institutes of Health (NIH) grant K23 AG055666 (to A.G.), the University of Kansas Medical Center Jared Grantham Kidney Institute Pilot Grant (to A.G.), NIH Clinical and Translational Science Award grant UL1 TR002366 (to the University of Kansas Medical Center), NIH grants P30 AG035982 (to the University of Kansas Alzheimer’s Disease Center), P30 DK106912 (to the University of Kansas Medical Center Jared Grantham Kidney Institute), R21 AG061549 (to E.D.V.) and S10 RR29577 (to W.M.B.), and a gift from Forrest and Sally Hoglund (to Hoglund Biomedical Imaging Center).

## Supplemental material Table of Contents

**Supplemental Table 1:** Transplantation related clinical characteristics of 22 end stage kidney disease (ESKD) patients who received kidney transplantation (KT).

**Supplemental Table 2:** Comparison of repeated MRI measurements in pre-KT patients. No changes were observed in one year.

**Supplementary Figure 1:** Scatterplots representing the Pearson’s correlation between **A)** CBF and eGFR, and **B)** CBF and serum creatinine 3 months post-KT. A strong correlation was not detected. CBF: cerebral blood flow, eGFR: estimated glomerular filtration rate, KT: kidney transplantation.

